# An improved and readily available version of *Bst* DNA Polymerase for LAMP, and applications to COVID-19 diagnostics

**DOI:** 10.1101/2020.10.02.20203356

**Authors:** Andre Maranhao, Sanchita Bhadra, Inyup Paik, David Walker, Andrew D. Ellington

## Abstract

Despite the fact that strand-displacing activity is of great utility for a variety of applications, including isothermal amplification assays, there are relatively few strand-displacing DNA polymerases. In particular, the thermotolerant DNA polymerase from *Geobacillus stearothermophilus (*previously *Bacillus stearothermophilus), Bst* DNA polymerase (*Bst* DNAP), is used in a variety of assays, including loop-mediated isothermal amplification. However, despite its wide use, its properties remain open to improvement, as has been demonstrated by a variety of engineering efforts, including the identification of point mutations that impact its robustness, strand-displacement capabilities, and nascent reverse transcriptase activity.

Interestingly, a strategy that has been commonly used to alter the capabilities of DNA polymerases, the addition of additional DNA- or RNA-binding domains, has yet to be applied to *Bst* DNAP. To this end, we now show that by adding fusion domains the performance of *Bst* DNAP in isothermal amplification assays, including its nascent RT activity, can be greatly improved. The impact of these improvements on the development of LAMP assays for the detection of SARS-CoV-2 is fully explored.

## Introduction

The development of isothermal amplification assays that are both sensitive and robust to sampling is key for continuing to mitigate the ongoing coronavirus pandemic (1). To this end, loop-mediated isothermal amplification has proven to be a useful assay for the detection of SARS-CoV-2 (2-5), including in clinical settings (6). However, LAMP is well-known to frequently produce spurious amplicons, even in the absence of template, and thus colorimetric and other methods that do not use sequence-specific probes may be at risk for generating false positive results (7), and we therefore developed oligonucleotide strand displacement probes, that are only triggered in the presence of specific amplicons. These probes are essentially the equivalent of TaqMan probes for qPCR, and can work either in an end-point or continuous fashion with LAMP (7). Base-pairing to the toehold region is extremely sensitive to mismatches, ensuring specificity, and the programmability of both primers and probes makes possible rapid adaptation to the evolution of new SARS-CoV-2 or other disease variants. We have also shown that higher order molecular information processing is also possible, such as integration of signals from multiple amplicons (8).

Our variant of LAMP, which we term LAMP-OSD (for Oligonucleotide Strand Displacement), is designed to be easy to use and interpret, and we have previously shown that it can sensitively and reliably detect SARS-CoV-2, including following direct dilution from saliva (8). Although we have largely mitigated non-specific signaling of LAMP and made it more robust for point of need application, the limited choice and supply, and concomitant expense of LAMP enzymes, constitutes a significant roadblock to widespread application of rapid LAMP-based diagnostics. We now demonstrate that the utility of LAMP-OSD can be further improved by utilizing new enzymes developed in our lab, and that can be made easily by any researcher.

## Results

### Engineering fusion variants of *Bst* DNAP

We centered our design around the large fragment of DNA polymerase I (Pol I) from *G. stearothermophilus* (*bst*, GenBank L42111.1), which is frequently used for isothermal amplification reactions (9-11) This fragment (hereafter Bst-LF) lacks a 310 amino acid N-terminal domain that is responsible for 5’ to 3’ exonuclease activity, leading to an increased efficiency of dNTP polymerization (12).

Many larger proteins suffer from slow or inefficient folding (13), and removal of the N-terminal portion may lead to increased protein degradation rates, decreased folding speed, and greater instability (14). Indeed, Bst-LF showed low yields upon purification. Therefore, in place of the exonuclease domain we sought to stabilize *Bst* DNAP via a fusion partner, the small F-actin binding protein villin, also referred to as the villin headpiece (15). The terminal thirty-five amino acids of the headpiece (HP35) consists of three α-helices that form a highly conserved hydrophobic core (16), exhibits co-translational, ultrafast, and autonomous folding properties that may circumvent kinetic traps during protein folding (17). The ultrafast folding property of the villin headpiece subdomain has made it a model for protein folding dynamics and simulation studies (18). In addition, HP35 displays thermostability with a transition midpoint (T_m_) of 70°C (16,17).

We extended HP35 by twelve amino acids to generate (HP47), where the additional amino acids serve to complete an additional alpha helix (N-3) in the structure, which further packs and stabilizes the hydrophobic core. The HP47 tag was added to the amino terminus of the large fragment of Bst-LF, leading to the enzyme we denote as Br512 (**Figure 1a**). Br512 also contains a N-terminal 8x His-tag for immobilized metal affinity chromatography (IMAC; Ni-NTA). The hypothesis that the villin headpiece could serve as an anchor to improve the folding and / or solubility of Bst-LF proved true, and purification of Br512 proved to be much better than for Bst-LF, ultimately yielding 35 mg of homogenous protein per liter (see also purification protocol, below).

**Figure 1.**
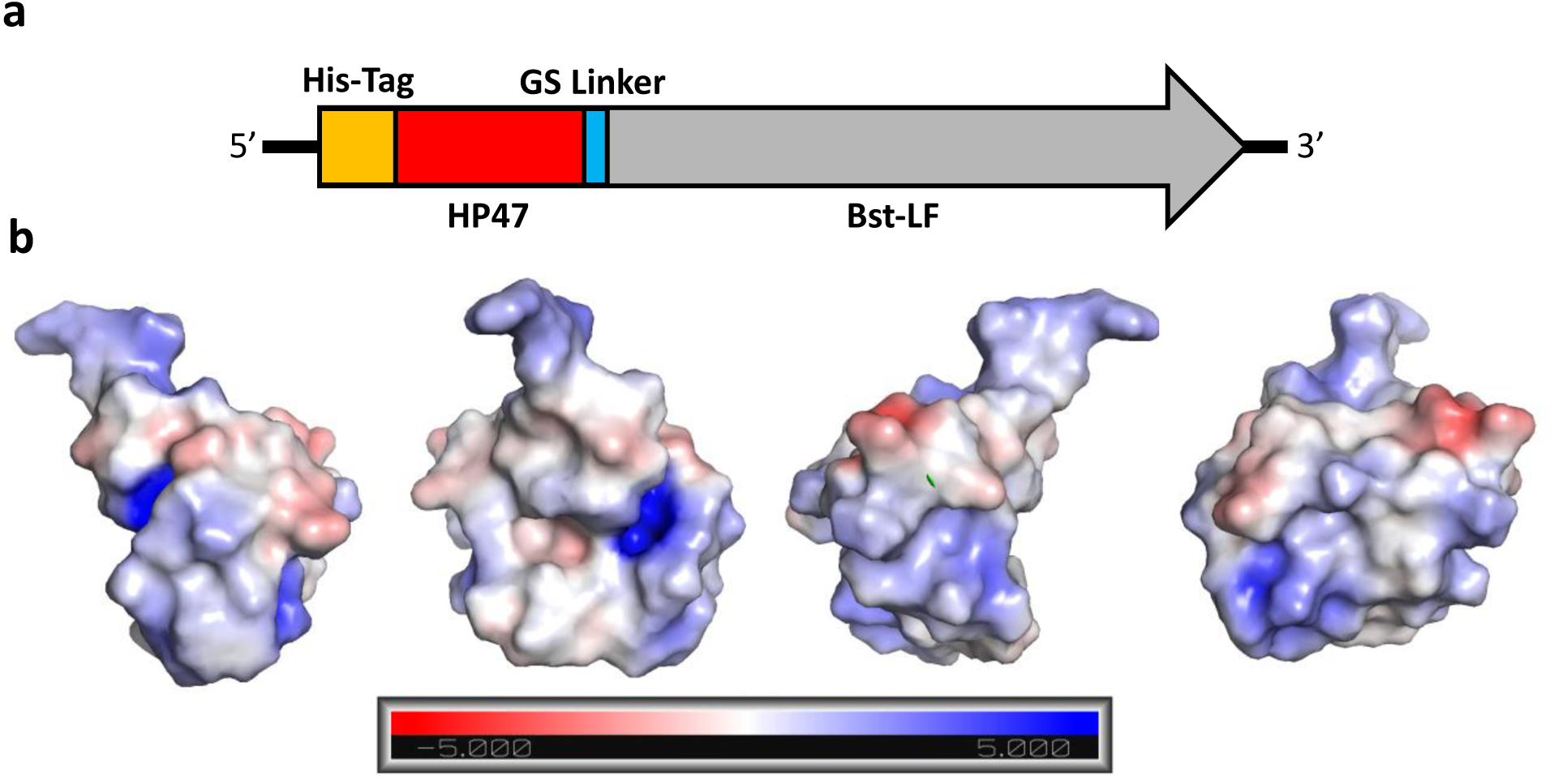
Graphical representation of Br512 and the electrostatic force map of HP47. (a) Br512 was constructed by fusing HP47 with a GS linker to the N-terminal of Bst-LF. A His-Tag was added at the N-terminal of the new fusion protein to aid with purification. (b) Models of HP47 electrostatic force using an Adaptive Poisson-Boltzmann Solver to identify surface charge. The charge designations are referenced in the bar at the bottom. Each graphic is the same model with different orientations rotated on the Y-Axis. Graphics were created in PyMol.

A cluster of positively charged amino acids are known to be crucial for the actin-binding activity of the headpiece domain (19), and thus another potential advantage of the use of HP47 is that it may allow better interactions with nucleic acid templates (17) (**Figure 1b**). In this regard, the HP47 fusion may act similarly to the DNA binding domains used in the construction of synthetic thermostable DNA polymerases that are commonly used for PCR. In the case of Phusion DNA polymerase, the addition of the Sso7d gene, a DNA binding protein from *Sulfolobus solfactaricus*, stabilizes the polymerase/DNA complex and enhances the processivity by up to 9 times, allowing longer amplicons in less time with less influence of PCR inhibitors (20). We further explored whether a similar improvement might prove true for isothermal amplification.

### Br512 performs comparably to Bst 2.0 with DNA templates

To assess whether the engineered changes introduced in Br512 had an impact on enzyme activity, we first compared the strand displacing DNA polymerase activity of Br512 with that of the wild type Bst-LF enzyme and a commercially-sourced engineered Bst-LF with improved amplification speed, Bst 2.0 (New England Biolabs). We set up duplicate LAMP-OSD assays (7) for the human glyceraldehyde 3-phosphate dehydrogenase (*gapd*) gene using either 16 units of Bst 2.0 (typical amount used in most of our LAMP-OSD assays), 20 picomoles (pm) of Bst-LF (a previously optimized amount), or 0.2 pm, 2 pm, 20 pm, or 200 pm of Br512. Real-time measurement of OSD probe fluorescence revealed that in the presence of 6000 template DNA copies the DNA polymerase activity of 20 pm of Br512 was comparable to that of 16 units of Bst 2.0 (**Supplementary Figure 1**). The addition of more Br512 did not yield further improvements, although lower amounts reduced the amplification efficiency. In the absence of specific templates, none of the enzymes generated false OSD signals.

When we set up assays with optimized enzyme amounts and either an optimized buffer system, G6 (developed for reverse transcription reactions with 4 mM (B) or 8 mM (D) Mg2^+^) (21), or Isothermal buffer (provided by New England Biolabs), we found that 20 pm of Br512 per LAMP-OSD assay performed comparably to 16 units of Bst 2.0 in terms of both speed and limit of detection (**Figure 2**). Bst-LF also demonstrated a similar detection limit but with a slower time to signal (**Figure 2**). Similar results were observed via real-time measurements of amplification kinetics using the fluorescent intercalating dye, EvaGreen in place of sequence-specific OSD probes (**Supplementary Figure 2**). Taken together, these results demonstrate that presence of the villin H47 fusion domain in Br512 improves its speed of amplification relative to Bst-LF bringing it at par with the DNA amplification speed of Bst 2.0.

**Figure 2.**
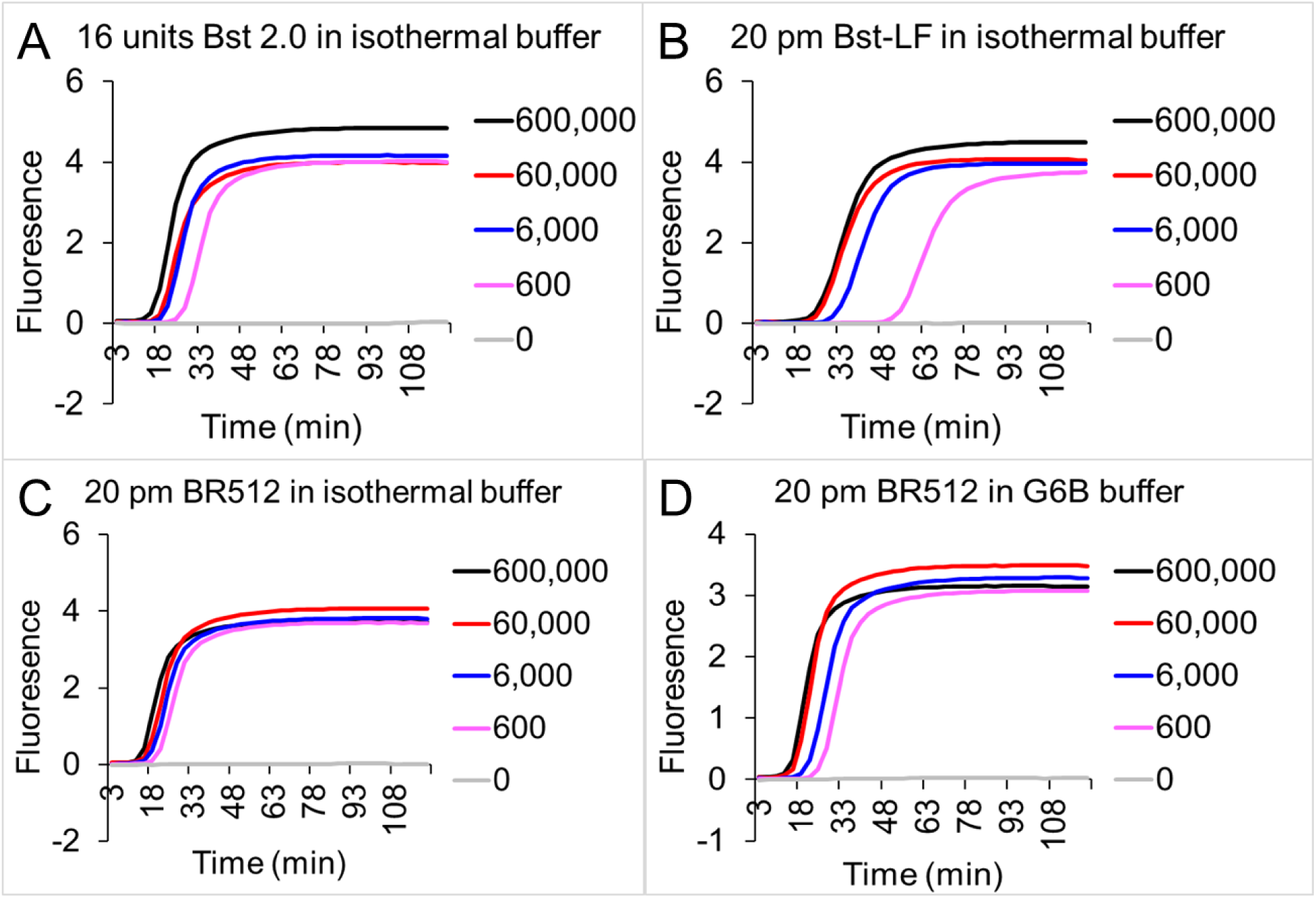
Comparison of Br512, Bst-LF, and Bst 2.0 in LAMP-OSD assays of DNA templates. LAMP-OSD assays for the human *gapd* gene were carried out with 16 units of commercially sourced Bst 2.0 (panel A), 20 pm of in-house purified Bst-LF (panel B), or 20 pm of Br512 (panels C and D) in the indicated reaction buffers. Amplification curves were observed in real-time at 65 °C by measuring OSD fluorescence in reactions seeded with 600,000 (black traces), 60,000 (red traces), 6,000 (blue traces), 600 (pink traces), and 0 (gray traces) copies of *gapd* plasmid templates.

### Br512 has superior performance in assays with RNA templates

Bst DNA polymerase has been described to possess an inherent reverse transcriptase (RT) activity with chemically diverse nucleic acid templates, including ribonucleic acid (RNA), α-l-threofuranosyl nucleic acid (TNA), and 2′-deoxy-2′-fluoro-β-d-arabino nucleic acid (FANA) (FANA), into DNA (22,23). Therefore, we tested the performance of Br512 in RT-LAMP-OSD assays in order to determine whether the engineered enzyme could be used for direct amplification from SARS-CoV-2 RNA. We set up duplicate reactions with three different primer sets that we had previously shown worked well with RT-LAMP-OSD (termed NB, Tholoth, and 6-Lamb (8)). These assays were seeded with 3000, 300, or 0 copies of the viral genomic RNA and endpoint OSD fluorescence was imaged following 60 min of amplification at 65 °C. All three RT-LAMP-OSD assays performed using Br512 developed bright green OSD fluorescence in the presence of viral genomic RNA indicating successful reverse transcription and LAMP amplification (**Figure 3**). While NB and 6-Lamb assays executed with Br512 could readily detect a few hundred viral genomes, the Tholoth assay produced visible signal only with a few thousand viral RNA copies. In contrast, Bst 2.0 demonstrated less robust reverse transcription ability, and failed to generate any OSD signal in the Tholoth assays, both in its companion Isothermal buffer (**Figures 3 and 4**) as well as in the G6D buffer (**Supplementary Figure 3**). Bst 2.0 could reverse transcribe and amplify NB and 6-Lamb sequences resulting in visible OSD fluorescence (**Figure 3**), but its detection limit for both assays was higher than with Br512. In all cases, in the absence of primers reactions remained dark.

**Figure 3.**
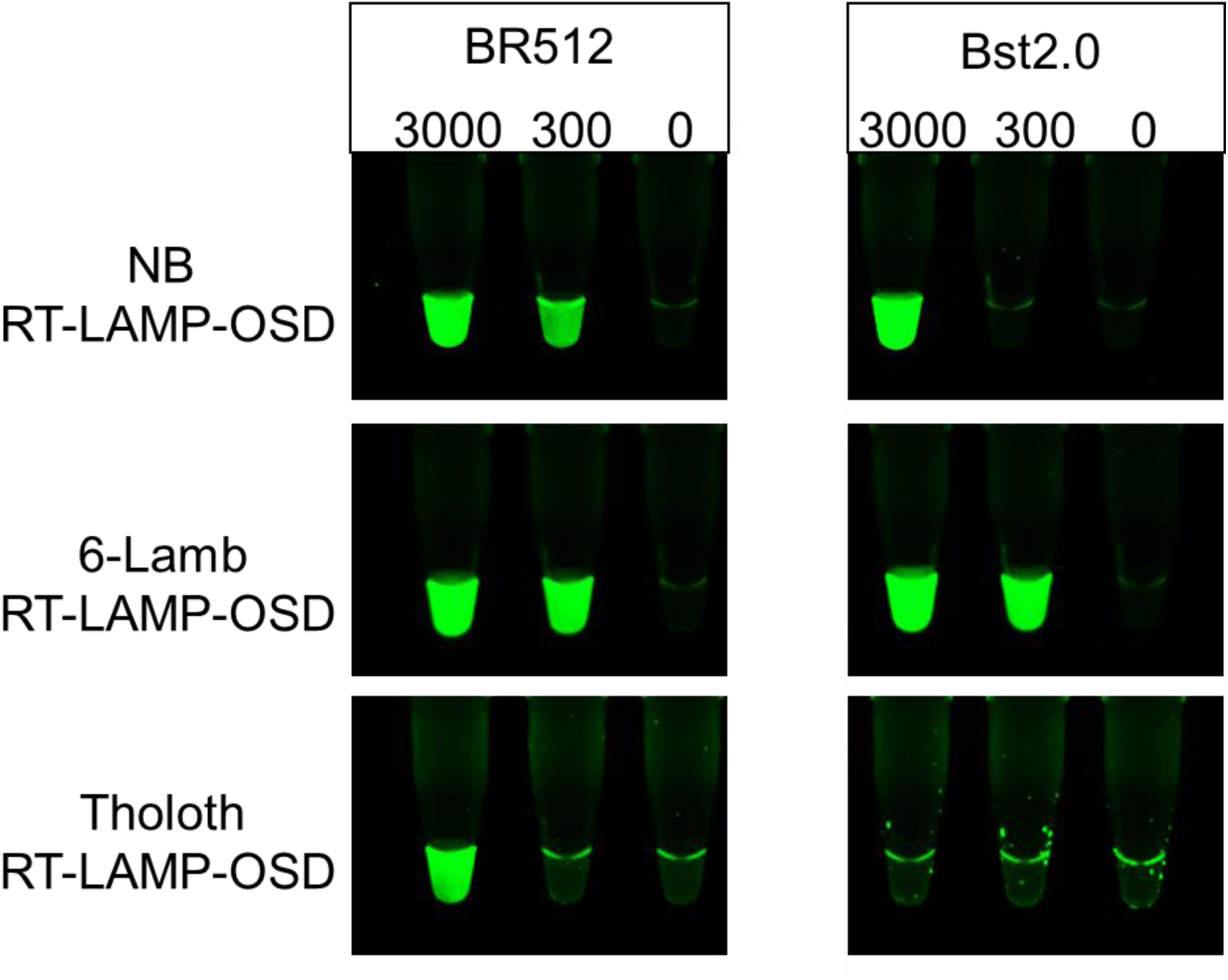
Comparison of Br512 and Bst 2.0 in visually-read RT-LAMP-OSD assays for SARS-CoV-2 viral genomic RNA. Three SARS-CoV-2-specific RT-LAMP-OSD assays, NB, 6-Lamb, and Tholoth, that target three different regions in the viral genomic RNA were operated using 20 pm of Br512 (left panels) in 1X G6D buffer or 16 units of commercially sourced Bst 2.0 (right panels) in 1X Isothermal buffer. Images of OSD fluorescence taken at assay endpoint (after 60 min of amplification at 65 °C followed by cooling to room temperature) are depicted.

**Figure 4.**
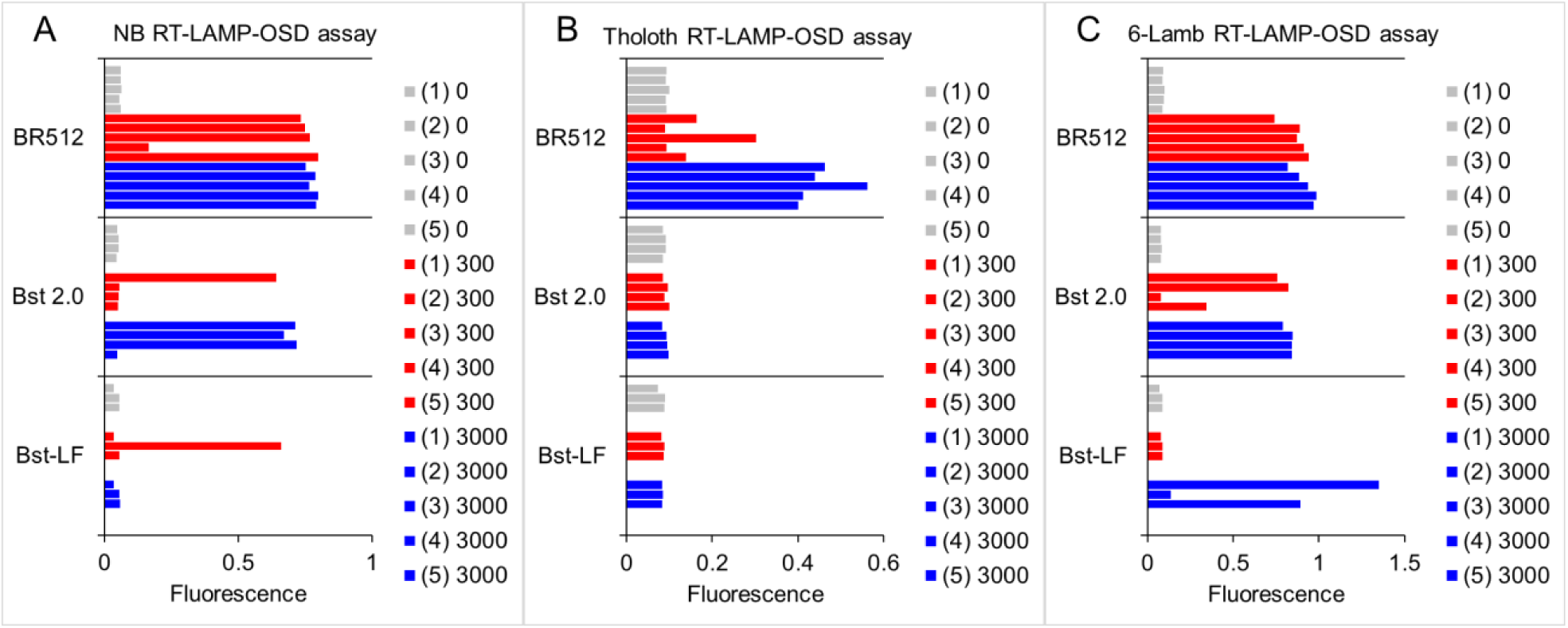
Comparison of Br512, Bst-LF, and Bst 2.0 in RT-LAMP-OSD assays for SARS-CoV-2 genomic RNA. Three SARS-CoV-2-specific RT-LAMP-OSD assays, NB (panel A), Tholoth (panel B), and 6-Lamb (panel C), were carried out with 20 pm of in-house purified Bst-LF, 16 units of commercially sourced Bst 2.0, or 20 pm of Br512 in G6D, isothermal, and G6D reaction buffers, respectively. OSD fluorescence measured at assay endpoint in reactions seeded with 3,000 (blue bars), 300 (red bars), or 0 (gray bars) copies of SARS-CoV-2 viral genomic RNA templates are depicted. Assay replicates in each panel are numbered 1 through 5. The real-time amplification kinetics of each reaction are detailed in **Supplementary Figure 4**.

The reliability of detection is an issue, especially at the limit of detection. Br512 could detect 300 SARS-CoV-2 genomes in 80% of NB and 100% of 6-Lamb assays, while Bst 2.0 was successful at detecting this copy number in 25% and 75% of the assays, respectively (**Figure 4**). In addition, Br512 generally demonstrated faster amplification kinetics compared to Bst 2.0 (**Supplementary Figure 4**). The superiority of Br512 as an enzyme for RT-LAMP was even more significant when compared to Bst-LF. The wild-type, parental enzyme failed to amplify the Tholoth RNA sequence, while only detecting SARS-CoV-2 genomic RNA in 16% of NB assays and 33% of 6-Lamb assays (**Figure 4**). Bst-LF also demonstrated slower amplification kinetics compared to both Br512 and Bst 2.0 (**Supplementary Figure 4**).

Having shown that individual SARS-CoV-2 amplicons could be successfully generated and detected by Br512 and RT-LAMP-OSD, we set up multiplex assays comprising primers and OSD probes for both the NB and 6-Lamb assays. As with RT-qPCR, such multiplex assays that detect multiple viral genes are of greatest utility for accurate confirmation of the virus (24,25). When irradiated SARS-CoV-2 virions were added to these assays, Br512 generated distinctly visible OSD signal from as few as 500 virions (**Supplementary Figure 5, panel A**). Duplicate assays executed using Bst 2.0 also produced bright OSD signals from 50,000 virions, while Bst 2.0 assays containing 5000 virions produced a dimmer OSD signal. 500 virions could not be directly detected using Bst 2.0 alone (**Supplementary Figure 5, panel B)**. The improved detection limit observed for Br512 might be due to the faster kinetics of amplification in multiplex RT-LAMP assays compared to Bst 2.0 (**Supplementary Figure 6**).

### Br512 can detect SARS-CoV-2 virions in saliva

LAMP is an appealing technology for rapid point-of-need testing because it does not require thermal cycling, and because the inhibitor tolerance of Bst DNA polymerase can enable direct analysis of clinical and environmental samples, thereby reducing assay complexity (26-28). Previously, we have described highly accurate detection of SARS-CoV-2 virions, including in saliva, using one-pot SARS-CoV-2 RT-LAMP-OSD assays via the standard commercial enzyme mix of Bst 2.0 and RTx (8).

To further determine whether Br512 can also function in crude clinical specimens we first seeded LAMP-OSD assays specific for human *gapd* sequence with human saliva. It should be emphasized that these reactions required no RNA preparation; they were direct dilution assays involving three microliters of saliva pre-heated at 95 °C for 10 min and 22 microliters of LAMP reaction mix. Following 60 min of amplification at 65 °C, both Br512 and Bst 2.0 generated bright OSD signals in the presence of saliva indicating successful amplification of endogenous *gapd* sequences (**Figure 5, panel A**). Assays lacking *gapd* LAMP primers remained as dark as control assays lacking saliva. These results suggest that Br512 is comparable to Bst 2.0 in its ability to amplify endogenous analyte sequences directly from saliva.

**Figure 5.**
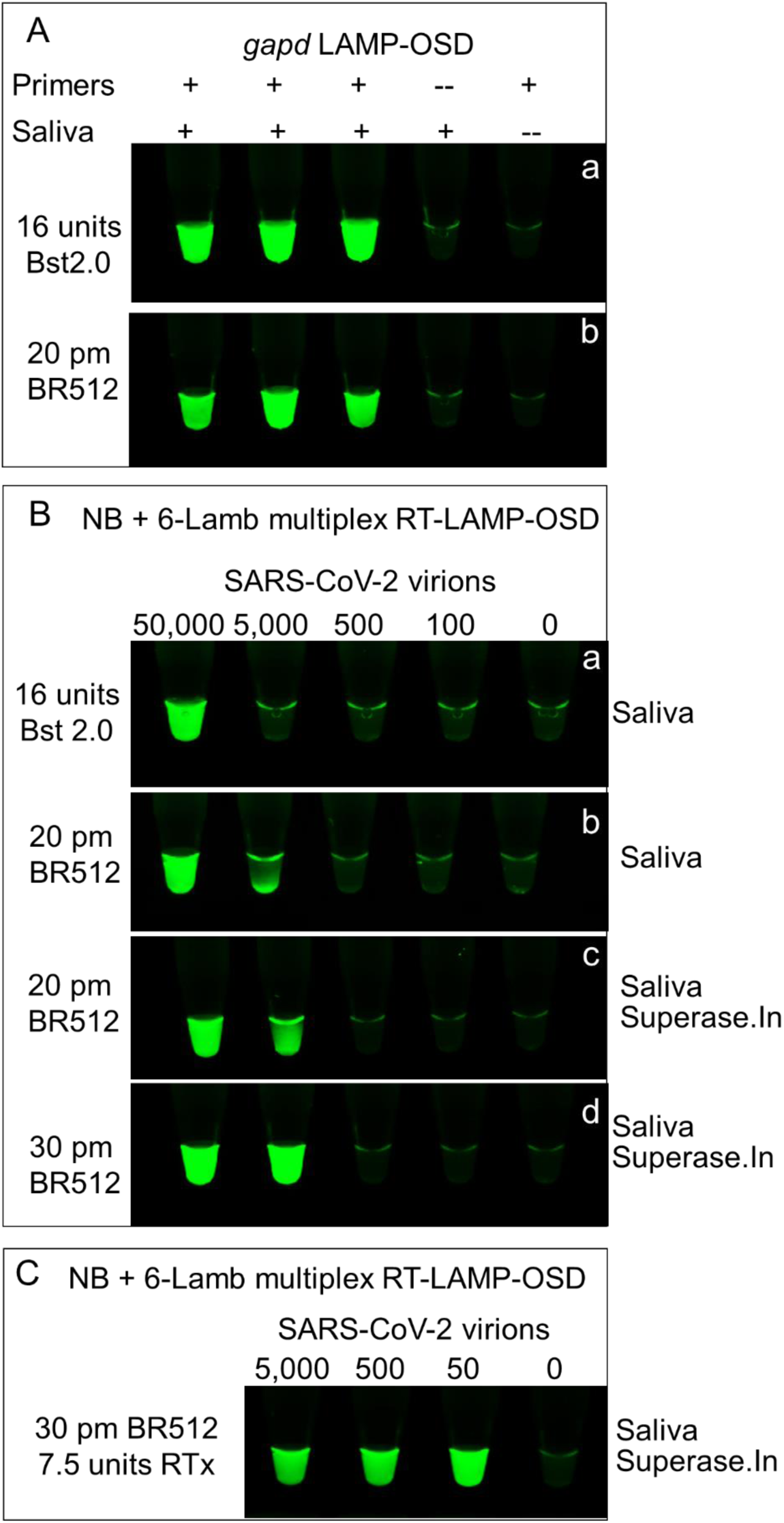
Comparison of Br512 and Bst2.0 *gapd* and SARS-CoV-2 assays in saliva. (A) Comparison of Br512 and Bst 2.0 DNA polymerase activity in LAMP-OSD assays of endogenous DNA templates in saliva. Duplicate LAMP-OSD assays with (+) or without (--) primers for the human *gapd* gene were executed either with 16 units of Bst 2.0 in isothermal buffer (NEB) or with 20 pm of Br512 in G6B buffer. Assays were seeded with 3 µL of water (--) or human saliva (+) heated for 10 min at 95 °C. Images of OSD fluorescence were taken at assay endpoint (after 60 min of amplification at 65 °C followed by cooling to room temperature). (B) Duplicate multiplex RT-LAMP-OSD assays containing primers and OSD probes for both NB and 6-Lamb SARS-CoV-2 assays were executed with the indicated amounts of either Bst 2.0 in Isothermal buffer (NEB) or Br512 in G6D buffer. Assays were seeded with the indicated number of copies of SARS-CoV-2 virions in the presence of 3 µL of human saliva that had been heated for 10 min at 95 °C. Some assays using Br512 also contained the RNase inhibitor, Superase.In. Images of OSD fluorescence were taken at assay endpoint after 60 min of amplification at 65 °C followed by cooling to room temperature. (C) Multiplex RT-LAMP-OSD assays containing primers and OSD probes for both the NB and 6-Lamb SARS-CoV-2 assays were executed using the indicated amounts of Br512 and RTx reverse transcriptase (NEB) enzymes in G6D buffer. Assays were seeded with the indicated number of copies of SARS-CoV-2 virions in the presence of 3 µL of heat treated human saliva. Images of OSD fluorescence were taken at assay endpoint after 60 min of amplification at 65 °C followed by cooling to room temperature.

When multiplex SARS-CoV-2 RT-LAMP-OSD assays were performed in the presence of 12% (v/v) human saliva in the reactions, both enzymes demonstrated reduced detection ability. In the presence of saliva, Br512 did not detect 500 virions and produced dimmer OSD signal from 5000 virions (**Figure 5, panel Ba**). In comparison, Bst 2.0 generated a signal only with 50,000 virions (**Figure 5, panel Bb**).

Inclusion of the RNase inhibitor, Superase.In, in the reaction overcame some of the inhibition observed relative to multiplex assays lacking saliva (**Supplementary Figure 5**) and allowed Br512 to produce brighter OSD signal from 5000 virions (**Figure 5, panel Bc**). Increasing the amount of Br512 in the reaction further boosted the signal (**Figure 5, panel Bd**).

These results demonstrate that it is feasible to conduct direct RNA detection in crude specimens using only Br512, albeit with a potential decline in detection limit. It is also conceivable that with further inhibition mitigation direct sample analysis with Br512 could approach levels seen with no-prep RT-qPCR with detection limits reported at 6-12 SARS-CoV-2 copies/µL (6000 – 12,000 copies/mL) with 5 µL sample volume per reaction for the SalivaDirect protocol implemented at Yale University (29) and 500 to 1000 viral particles/mL with 10 µL sample volume per reaction for the sample prep protocol developed at University of Illinois (30). Furthermore, by including a reverse transcriptase (RTx; NEB) along with Br512 we were able to reduce the limit of detection for direct saliva analysis to 50 viral copies per reaction, starting from 3 µL saliva input volume (**Figure 5, panel C**).

### Br512 is robust to different reaction preparations and conditions

Ready-to-use, freeze-dried assay mixes facilitate large scale distribution and implementation of rapid assays. To determine whether Br512-based single enzyme RT-LAMP-OSD assays might be amenable to drying, we prepared master mixes for the SARS-CoV-2 multiplex assay containing both NB and 6-Lamb primers and OSD probes, along with either 20 pm or 30 pm of Br512 and subjected them to lyophilization. After two days we rehydrated the dry assay pellets and seeded them with different amounts of SARS-CoV-2 viral genomic RNA. As shown in **Figure 6**, both the 20 pm and 30 pm lyophilized BR512 assays produced visible OSD fluorescence in the presence of as few as 300 viral genomic RNA. Assays lyophilized with more enzyme were generally brighter. These results demonstrate that Br512 is amenable to production of freeze dried single enzyme RT-LAMP assay mixes that are ready for diagnostics simply upon rehydration.

**Figure 6.**
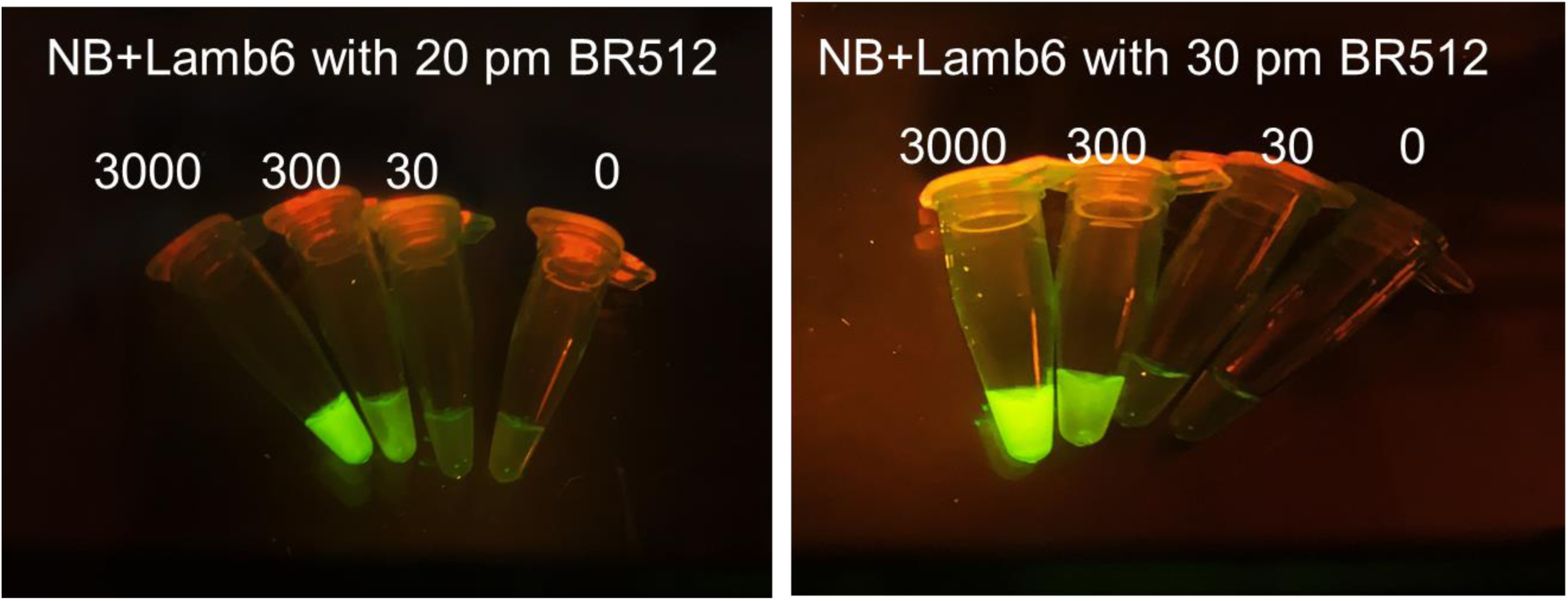
Assessment of lyophilized Br512 multiplex SARS-CoV-2 RT-LAMP-OSD assays. Lyophilized multiplex RT-LAMP-OSD assays prepared with 20 or 30 picomoles of glycerol-free Br512 enzymes, primers, and OSD probes for both NB and 6-Lamb assays were tested with the indicated number of copies of SARS-CoV-2 genomic RNA. Images of OSD fluorescence were taken after 60 min of amplification at 65 °C followed by cooling to room temperature.

Ideally, diagnostic enzymes should be robust to some variance in reaction conditions, especially those that may be inadvertently introduced by user error or sample variation. We analyzed the effect of varying buffer and salt concentrations on the activity of Br512 with *gapd* LAMP-OSD reactions. Compared to the optimized G6 buffer previously described, a 66% drop in Tris buffer concentration, 100% decrease or 150% increase in (NH)_2_SO_4_ amount, or 75% decrease in KCl did not cause significant perturbations in the amplification speed or detection limit of Br512 (**Supplementary Figure 7**). These results suggest that Br512 can perform robustly under varied reaction conditions.

### Distribution of enzyme

Constructs for the production of enzyme can be obtained from Addgene, or via requests to the Ellington lab (send requests to, with a cc: to). Enzyme can be purified via the methods described in **Materials and Methods**.

## Materials and Methods

### Chemicals and reagents

All chemicals were of analytical grade and were purchased from Sigma-Aldrich (St. Louis, MO, USA) unless otherwise indicated. All commercially sourced enzymes and related buffers were purchased from New England Biolabs (NEB, Ipswich, MA, USA) unless otherwise indicated. All oligonucleotides and gene blocks (**Supplementary Table 1**) were obtained from Integrated DNA Technologies (IDT, Coralville, IA, USA). SARS-CoV-2 genomic RNA and inactivated virions were obtained from American Type Culture Collection, Manassas, VA, USA. A single sample of human saliva was collected from a lab member by passive drooling after a 4 h period without eating or drinking. The saliva sample was stored at -20 °C until use.

### Br512 purification protocol

The overall scheme for Br512 purification is shown in **Supplementary Figure 8**. In short, Br512 was cloned into an in-house *E. coli* expression vector under the control of a T7 RNA polymerase promoter (pKAR2). Full sequence and annotations of the pKAR2-Br512 plasmid are available in **Supplementary Table 2**. The Br512 expression construct pKAR2-Br512 was then transformed into *E. coli* BL21(DE3) (NEB, C2527H). A single colony was seed cultured overnight in 5 mL of superior broth (Athena Enzyme Systems, 0105). The next day, 1 mL of seed culture was inoculated into 1 L of superior broth and grown at 37 ^°^C until it reached an OD600 of 0.5-0.6. Enzyme expression was induced with 1 mM IPTG and 100 ng/mL of anhydrous tetracycline (aTc) at 18 ^°^C for 18 h (or overnight). The induced cells were pelleted at 5000xg for 10 min at 4^°^C and resuspended in 30mL of ice-cold lysis buffer (50 mM Phosphate Buffer, pH 7.5, 300 mM NaCl, 20 mM imidazole, 0.1% Igepal CO-630, 5 mM MgSO4, 1 mg/mL HEW Lysozyme, 1x EDTA-free protease inhibitor tablet, Thermo Scientific, A32965). The samples were then sonicated (1 sec ON, 4 sec OFF) for a total time of 4 minutes with 40% amplitude. The lysate was centrifuged at 35,000 xg for 30 minutes at 4 °C. The supernatant was transferred to a clean tube and filtered through a 0.2 μm filter.

Protein from the supernatant was purified using metal affinity chromatography on a Ni-NTA column. Briefly, 1mL of Ni-NTA agarose slurry was packed into a 10mL disposable column and equilibrated with 20 column volume (CV) of equilibration buffer (50 mM Phosphate Buffer, pH 7.5, 300 mM NaCl, 20 mM imidazole). The sample lysate was loaded onto the column and the column was developed by gravity flow. Following loading, the column was washed with 20 CV of equilibration buffer and 5 CV of wash buffer (50 mM Phosphate Buffer, pH 7.5, 300 mM NaCl, 50 mM imidazole). Br512 was eluted with 5 mL of elution buffer (50 mM Phosphate Buffer, pH 7.5, 300 mM NaCl, 250 mM imidazole). The eluate was dialyzed twice with 2L of Ni-NTA dialysis buffer (40 mM Tris-HCl, pH 7.5, 100 mM NaCl, 1 mM DTT, 0.1% Igepal CO-630). The dialyzed eluate was further passed through an equilibrated 5 mL heparin column (HiTrap^TM^ Heparin HP) on a FPLC (AKTA pure, GE healthcare) and eluted using a linear NaCl gradient generated from heparin buffers A and B (40 mM Tris-HCl, pH 7.5, 100 mM NaCl for buffer A; 2M NaCl for buffer B, 0.1% Igepal CO-630). The collected final eluate was dialyzed first with 2 L of heparin dialysis buffer (50 mM Tris-HCl, pH 8.0, 50 mM KCl, 0.1% Tween-20) and second with 2 L of final dialysis buffer (50% Glycerol, 50 mM Tris-HCl, pH 8.0, 50 mM KCl, 0.1% Tween-20, 0.1% Igepal CO-630, 1 mM DTT). The purified Br512 was quantified by Bradford assay and SDS-PAGE/coomassie gel staining alongside a bovine serum albumin (BSA) standard.

### Real-time *gapd* LAMP-OSD

LAMP-OSD reaction mixtures were prepared in 25 µL volume containing indicated amounts of human glyceraldehyde-3-phosphate dehydrogenase (*gapd*) DNA templates along with a final concentration of 1.6 μM each of BIP and FIP primers, 0.4 μM each of B3 and F3 primers, and 0.8 μM of the loop primer. Amplification was performed in one of the following buffers – 1X Isothermal buffer (NEB) (20 mM Tris-HCl, 10 mM (NH4)_2_SO_4_, 10 mM KCl, 2 mM MgSO_4_, 0.1% Triton X-100, pH 8.8), 1X G6B buffer (60 mM Tris-HCl, pH 8.0, 2 mM (NH_4_)_2_SO_4_, 40 mM KCl, 4 mM MgCl_2_), 1X G1B buffer (60 mM Tris-HCl, pH 8.0, 5 mM (NH_4_)_2_SO4, 10 mM KCl, 4 mM MgSO_4_, 0.01% Triton X-100), 1X G2A buffer (20 mM Tris-HCl, pH 8.0, 5 mM (NH_4_)_2_SO_4_, 10 mM KCl, 4 mM MgCl_2_, 0.01% Triton X-100), or 1X G3A buffer (60 mM Tris-HCl, pH 8.0, 10 mM KCl, 4 mM MgCl_2_, 0.01% Triton X-100). The buffer was appended with 1 M betaine, 0.4 mM dNTPs, 2 mM additional MgSO_4_ (only for reactions in Isothermal buffer), and either Bst 2.0 DNA polymerase (16 units), Bst-LF DNA polymerase (20 pm), or Br512 DNA polymerase (0.2 pm, 2 pm, 20 pm, or 200 pm). Assays read using OSD probes received 100 nM of OSD reporter prepared by annealing 100 nM fluorophore-labeled OSD strands with a 5-fold excess of the quencher-labeled OSD strands by incubation at 95 °C for 1 min followed by cooling at the rate of 0.1 °C/sec to 25 °C. Assays read using intercalating dyes received 1X EvaGreen (Biotium, Freemont, CA, USA) instead of OSD probes. For real-time signal measurement these LAMP reactions were transferred into a 96-well PCR plate, which was incubated in a LightCycler 96 real-time PCR machine (Roche, Basel, Switzerland) maintained at 65 °C for 90 min. Fluorescence signals were recorded every 3 min in the FAM channel and analyzed using the LightCycler 96 software. For assays read using EvaGreen, amplification was followed by a melt curve analysis on the LightCycler 96 to distinguish target amplicons from spurious background.

### Endpoint *gapd* LAMP-OSD

LAMP-OSD reaction mixtures for visual endpoint readout of OSD fluorescence were prepared in 25 µL volume containing either 1X Isothermal buffer and 16 units of Bst 2.0 or 1X G6B buffer and 20 pm of Br512. The reactions also contained 1 M betaine, 1.4 mM dNTPs, 2 mM additional MgSO_4_ (only for reactions in Isothermal buffer), and 100 nM OSD reporter prepared by annealing 100 nM fluorophore-labeled OSD strands with a 2-fold excess of the quencher-labeled OSD strands by incubation at 95 °C for 1 min followed by cooling at the rate of 0.1 °C/sec to 25 °C. A final concentration of 1.6 μM each of BIP and FIP primers, 0.4 μM each of B3 and F3 primers, and 0.8 μM of the loop primer were added to some reactions while control assays without primers received the same volume of water. Some assays were seeded with 3 µL of human saliva that had been heated at 95 °C for 10 min while other assays received the same volume of water. All assays were incubated in a thermocycler maintained at 65 °C for 60 min following which endpoint OSD fluorescence was imaged using a ChemiDoc camera (Bio-Rad Laboratories, Hercules, CA, USA).

### SARS-CoV-2 RT-LAMP-OSD assays

Individual 25 µL RT-LAMP-OSD assays were assembled either in 1X Isothermal buffer containing 16 units of Bst 2.0 or in 1X G6D buffer (60 mM Tris-HCl, pH 8.0, 2 mm (NH_4_)_2_SO_4_, 40 mM KCl, 8 mM MgCl_2_) containing 20 pm of Br512, 20 pm of Bst-LF, or 16 units of Bst 2.0. The buffer was supplemented with 1.4 mM dNTPs, 0.4 M betaine, 6 mM additional MgSO_4_ (only for reactions in Isothermal buffer), and 2.4 µM each of FIP and BIP, 1.2 µM of indicated loop primers, and 0.6 µM each of F3 and B3 primers. Amplicon accumulation was measured by adding OSD probes. First, Tholoth, Lamb, and NB OSD probes were prepared by annealing 1 µM of the fluorophore-labeled OSD strand with 2 µM, 3 µM, and 5 µM, respectively of the quencher-labeled strand in 1X Isothermal buffer. Annealing was performed by denaturing the oligonucleotide mix at 95 °C for 1 min followed by slow cooling at the rate of 0.1 °C/s to 25 °C. Excess annealed probes were stored at -20 °C. Annealed Tholoth, Lamb, and NB OSD probes were added to their respective RT-LAMP reactions at a final concentration of 100 nM of the fluorophore-bearing strand. The assays were seeded with indicated amounts of SARS-CoV-2 viral genomic RNA in TE buffer (10 mM Tris-HCl, pH 7.5, 0.1 mM EDTA) and either incubated for 1 h in a thermocycler maintained at 65 °C for endpoint readout or transferred to a 96-well plate and incubated in the LightCycler 96 real-time PCR machine maintained at 65 °C for real time measurement of amplification kinetics. Endpoint OSD fluorescence was read visually using a blue LED torch with orange filter and imaged using a cellphone camera or a ChemiDoc (Bio-Rad) camera. OSD fluorescence in assays incubated in the real-time PCR machine was measured every 3 min in the FAM channel and analyzed using the LightCycler 96 software.

Multiplex RT-LAMP-OSD assays comprising 6-Lamb and NB primers and OSD probes were set up using the same conditions as above except that the total LAMP primer amounts were made up of equimolar amounts of 6-Lamb and NB primers supplemented with 0.2 µM each of additional NB FIP and BIP primers. Some multiplex assays received 30 pm Br512 instead of 20 pm enzyme and others received 7.5 units of RTx reverse transcriptase (NEB) along with 30 pm of Br512. Some multiplex assays also received 20 units of the RNase inhibitor, SUPERase.In (Thermo Fisher Scientific, Waltham, MA). Multiplex assays were seeded with indicated amounts of either SARS-CoV-2 viral genomic RNA or inactivated virions in the presence of either 3 µL of TE buffer or human saliva pre-heated at 95 °C for 10 min. The assays were either incubated for 1 h in a thermocycler maintained at 65 °C for endpoint readout or transferred to a 96-well plate and incubated in the LightCycler 96 real-time PCR machine maintained at 65 °C for real time measurement of amplification kinetics. Endpoint OSD fluorescence was read visually using a blue LED torch with orange filter and imaged using a cellphone camera or a ChemiDoc (Bio-Rad) camera. OSD fluorescence in assays incubated in the real-time PCR machine was measured every 3 min in the FAM channel and analyzed using the LightCycler 96 software.

### Lyophilization of Br512

Multiplex SARS-CoV-2 assay reagent mixes were prepared by combining dNTPs, NB and 6-Lamb primers and OSD probes, trehalose, and glycerol-free Br512 in the following amounts per individual reaction – (i) 35 nanomoles of dNTPs, (ii) 30 pm each of 6-Lamb FIP and BIP, 15 pm each of 6-Lamb LF and LB loop primers, 7.5 pm each of 6-Lamb F3 and B3 primers, (iii) 35 pm each of NB FIP and BIP, 15 pm of NB LB loop primer, 7.5 pm each of NB F3 and B3 primers, (iv) 2.5 pm of NB fluorophore labeled OSD strand pre-annealed with 5-fold excess of the quencher labeled OSD strand, (v) 2.5 pm of Lamb fluorophore labeled OSD strand pre-annealed with a 3-fold excess of the quencher labeled OSD strand, (vi) 1.25 micromoles of trehalose, and (vii) 20 pm or 30 pm of BR512. The reagent mixes were distributed in 0.2 mL PCR tubes and frozen for 1 h on dry ice prior to lyophilization for 2.5 h at 197 mTorr and -108 °C using the automated settings in a VirTis Benchtop Pro lyophilizer (SP Scientific, Warminster, PA, USA). Lyophilized assays were stored with desiccant (Drierite) at -20 °C until use.

Lyophilized assays were rehydrated immediately prior to use by adding 22 µL of 1X G6D buffer containing 10 micromoles of betaine. Rehydrated assays were seeded with indicated amounts of SARS-CoV-2 viral genomic RNA in a total volume of 3 µL and incubated for 1 h in a thermocycler maintained at 65 °C. Endpoint OSD fluorescence was read visually using a blue LED torch with orange filter and imaged using a cellphone camera.

## Discussion

While previously we had demonstrated the utility and robustness of LAMP-OSD for SARS-CoV-2 detection, including in potential point-of-care formats, the scaled implementation of these solutions required further innovations, especially for low-resource settings that may not have access to commercial enzymes, reaction mixes, or kits, and that therefore may require local production of the reagents. In this regard, the development of Br512 provides a potential alternative solution for assay development. The use of the villin headpiece as a fusion tag for the strand-displacing *Bst* DNA polymerase serves two purposes: one to stabilize the protein for purification and eventually for use in assays that require little or no sample preparation; and two, to assist with interactions with nucleic acids, due to the “Janus” charged nature of the headpiece, where one side is primarily positively charged and the opposite side negatively charged. In this regard, it functions much like Sso7d and similar nucleic acid-binding domains that have previously been appended to polymerases (e.g., the well-known and widely used Phusion DNA polymerase), but with additional advantages. In the current instance, we suspect (but have yet to fully prove) that the addition of the villin headpiece leads to more robust interactions with RNA, and increased reverse transcriptase activity. Even so, it is recommended that Br512 be used with a dedicated reverse transcriptase (herein we use RTx, from New England Biolabs) to achieve the highest sensitivity for the detection of SARS-CoV-2.

## Conclusion

Given the prevalence of isothermal amplification assays for diagnostics, especially for point-of-care screening, the development of a Bst 2.0 alternative with superior properties is of potential utility. Br512 can be readily prepared, and gives similar or better limits of detection than Bst 2.0 in a variety of conditions, including in multiplex assays for the direct detection of inactivated viral particles in saliva. Coupling Br512 with a reverse transcriptase further improves its capabilities, and can provide sensitivities for detection that rival those of direct dilution RT-qPCR.

## Data Availability

All data pertaining to the study have been provided in the manuscript.

