## Supplementary information for "An improved and readily available version of *Bst* DNA Polymerase for LAMP, and applications to COVID-19 diagnostics"

Andre Maranhao[1], Sanchita Bhadra[1], Inyup Paik, David Walker, and Andrew D. Ellington[2]
Center for Systems and Synthetic Biology
Department of Molecular Biosciences
University of Texas at Austin
Austin, TX

Supplementary information

Supplementary Figure 1

Supplementary Figure 2

Supplementary Figure 3

Supplementary Figure 4

Supplementary Figure 5

Supplementary Figure 6

Supplementary Figure 7

Supplementary Figure 8

Supplementary Table 1

Supplementary Table 2

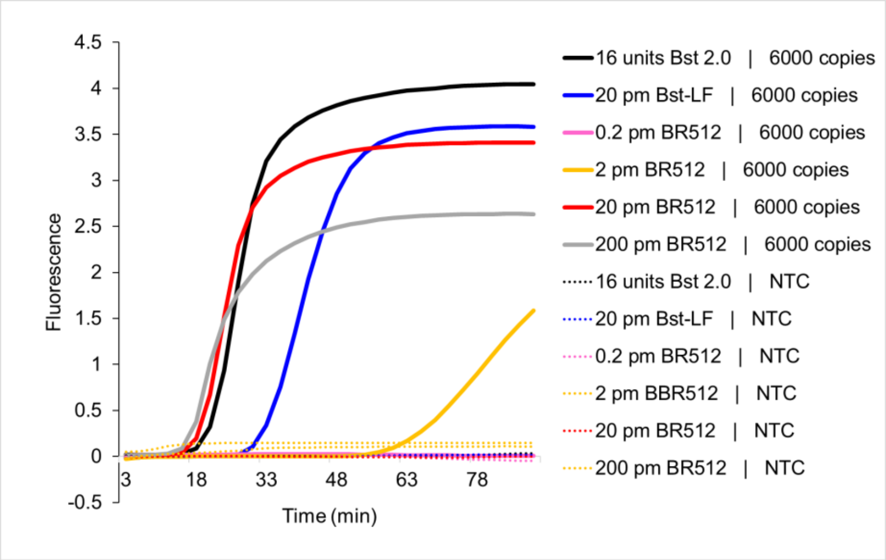


**Supplementary Figure 1. Effect of varying amounts of Br512 on LAMP-OSD of DNA templates.** Indicated amounts of Br512 were compared with indicated amounts of in-house purified Bst-LF and commercially sourced Bst 2.0 in human *gapd* gene-specific LAMP-OSD assays operated in 1X isothermal buffer (NEB). Reactions were seeded with either 6000 copies of *gapd* plasmid template or with no specific templates (NTC). Amplification curves generated by real-time measurement of OSD fluorescence at 65 °C are depicted.

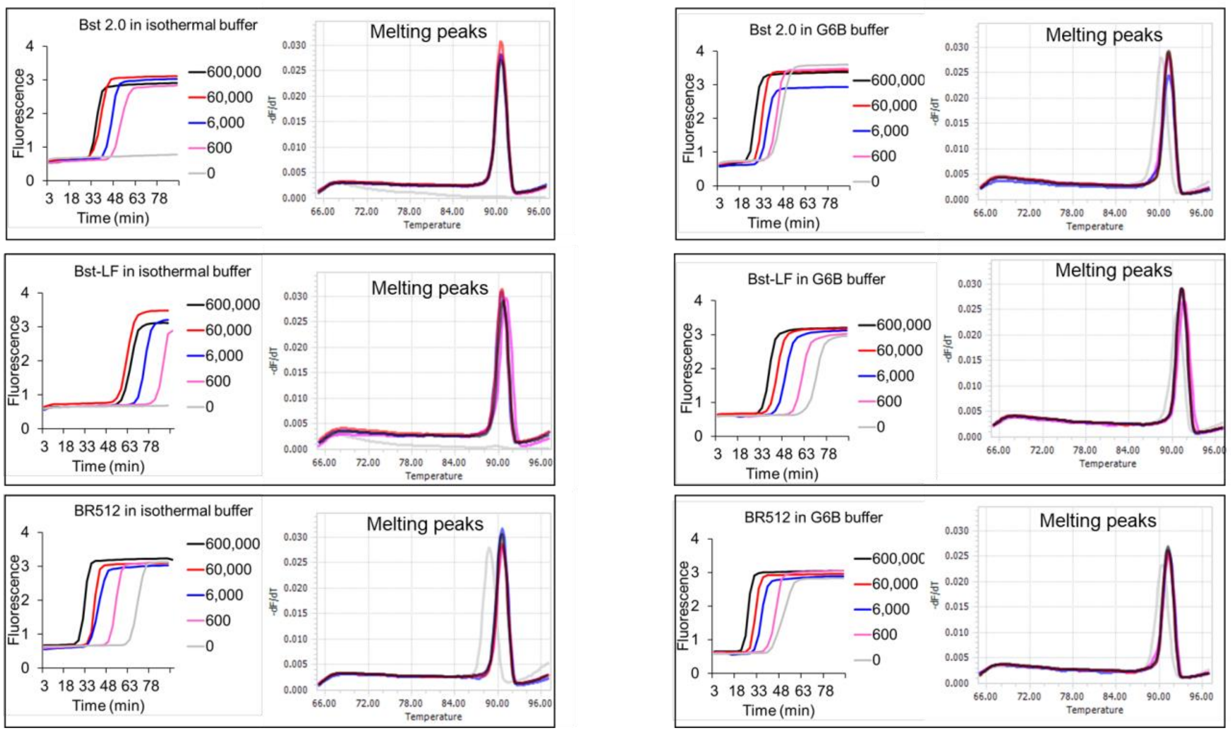


**Supplementary Figure 2. Comparison of Br512, Bst-LF, and Bst 2.0 in LAMP assays of DNA templates read using EvaGreen intercalating dye.** LAMP assays for human *gapd* gene were operated using Bst 2.0, Bst-LF, or Br512 in indicated reaction buffers. Amplification curves observed in real-time at 65 °C by measuring EvaGreen fluorescence in reactions seeded with 600,000 (black traces), 60,000 (red traces), 6,000 (blue traces), 600 (pink traces), and 0 (gray traces) copies of *gapd* plasmid templates are depicted. LAMP amplicons were analyzed using the ‘melt curve analysis’ on LightCycler 96 real-time PCR machine and resulting melting peaks are indicated in the corresponding colored traces.

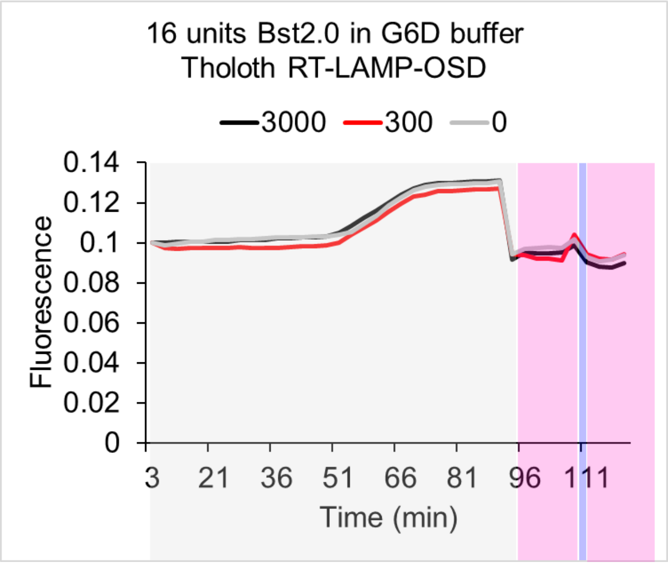


**Supplementary Figure 3. Bst 2.0 Tholoth RT-LAMP-OSD assay executed in G6D buffer.** Tholoth RT-LAMP-OSD assays for SARS-CoV-2 were operated using Bst 2.0 in G6D reaction buffer. OSD fluorescence measured in real-time during assay incubation at 65 °C are depicted within gray shaded boxes for reactions seeded with 3,000 (black traces), 300 (red traces), or 0 (gray traces) copies of SARS-CoV-2 genomic RNA templates. Post-amplification phase OSD signal measured at 37 °C before and after a 1 min DNA denaturation step at 95 °C (in blue shaded region) are depicted within the pink shaded regions.

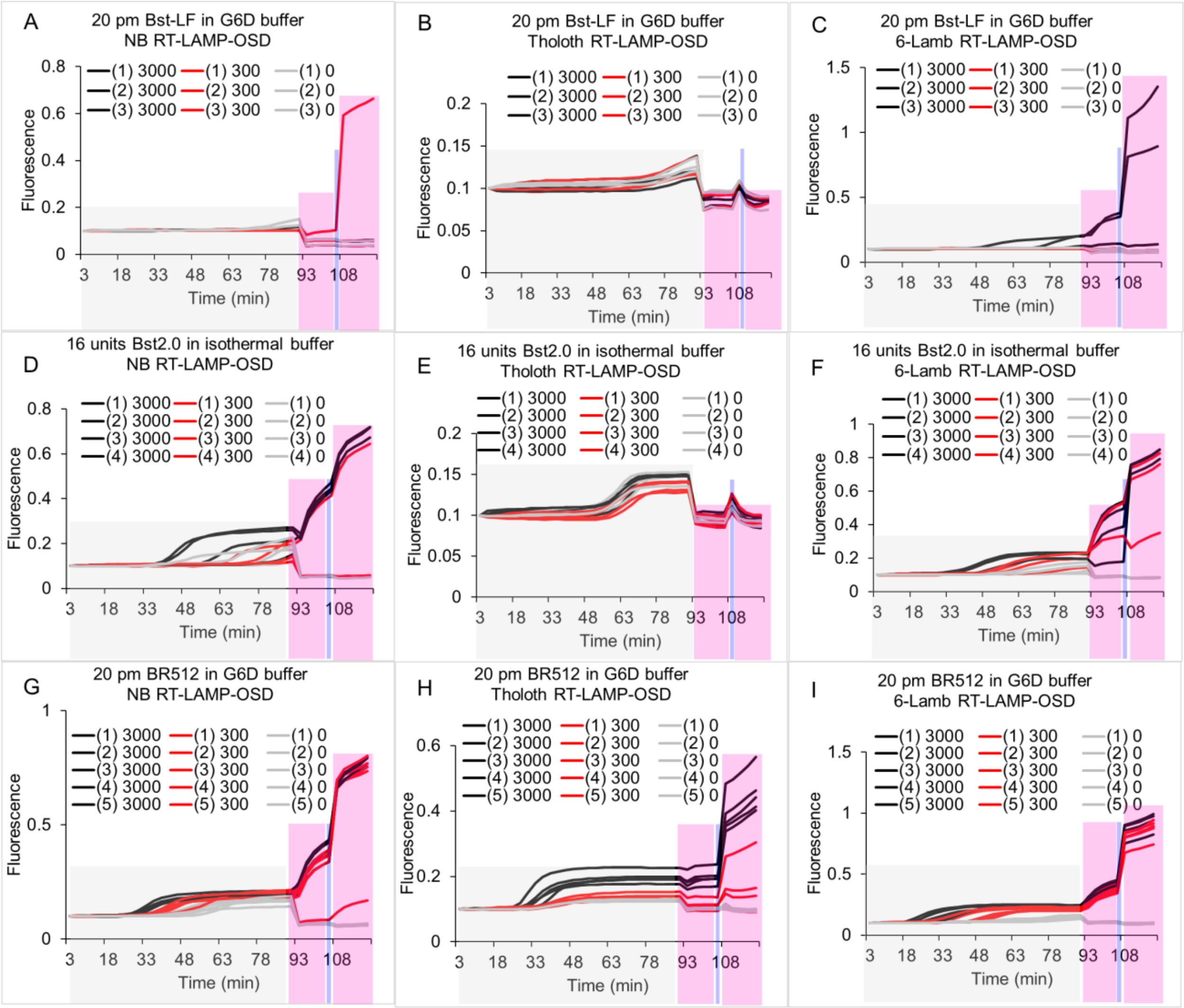


**Supplementary Figure 4. Comparison of Br512, Bst-LF, and Bst 2.0 in RT-LAMP-OSD assays for SARS-CoV-2 genomic RNA.** Three SARS-CoV-2-specific RT-LAMP-OSD assays, NB (panels A, D, and G), Tholoth (panels B, E, and H), and 6-Lamb (panels C, F, and I), were operated using 20 pm of in-house purified Bst-LF (panels A, B, and C), 16 units of commercially sourced Bst 2.0 (panels D, E, and F), or 20 pm of Br512 (panels G, H, and I) in indicated reaction buffers. Amplification kinetics at 65 °C observed in real-time by measuring OSD fluorescence in reactions seeded with 3,000 (black traces), 300 (red traces), or 0 (gray traces) copies of SARS-CoV-2 viral genomic RNA templates are depicted within gray shaded boxes. Post-amplification OSD signal measured at 37 °C before and after a 1 min DNA denaturation step at 95 °C (in blue shaded region) are depicted within the pink shaded regions. Assay replicates in each panel are numbered 1 through 5.

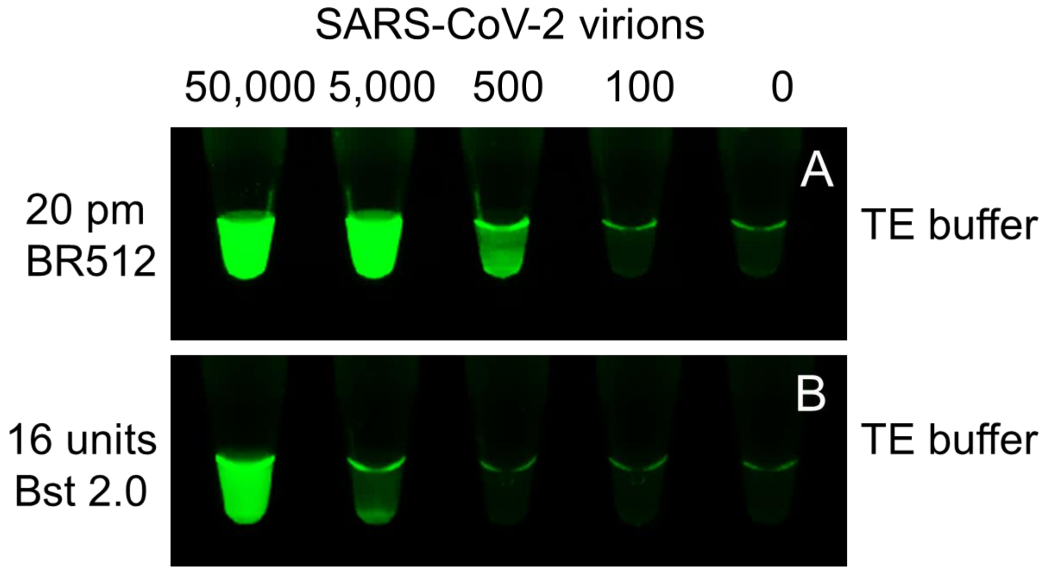


**Supplementary Figure 5. Comparison of Br512 and Bst2.0 SARS-CoV-2 RT-LAMP-OSD assays using SARS-CoV-2 virions.** Duplicate multiplex RT-LAMP-OSD assays containing primers and OSD probes for both NB and 6-Lamb SARS-CoV-2 assays were executed with indicated amounts of either Bst 2.0 in isothermal buffer (NEB) or Br512 in G6D buffer. Assays were seeded with indicated copies of SARS-CoV-2 virions in the presence of 3 µL of TE buffer. Images of OSD fluorescence taken at assay endpoint after 60 min of amplification at 65 °C followed by cooling to room temperature are depicted.

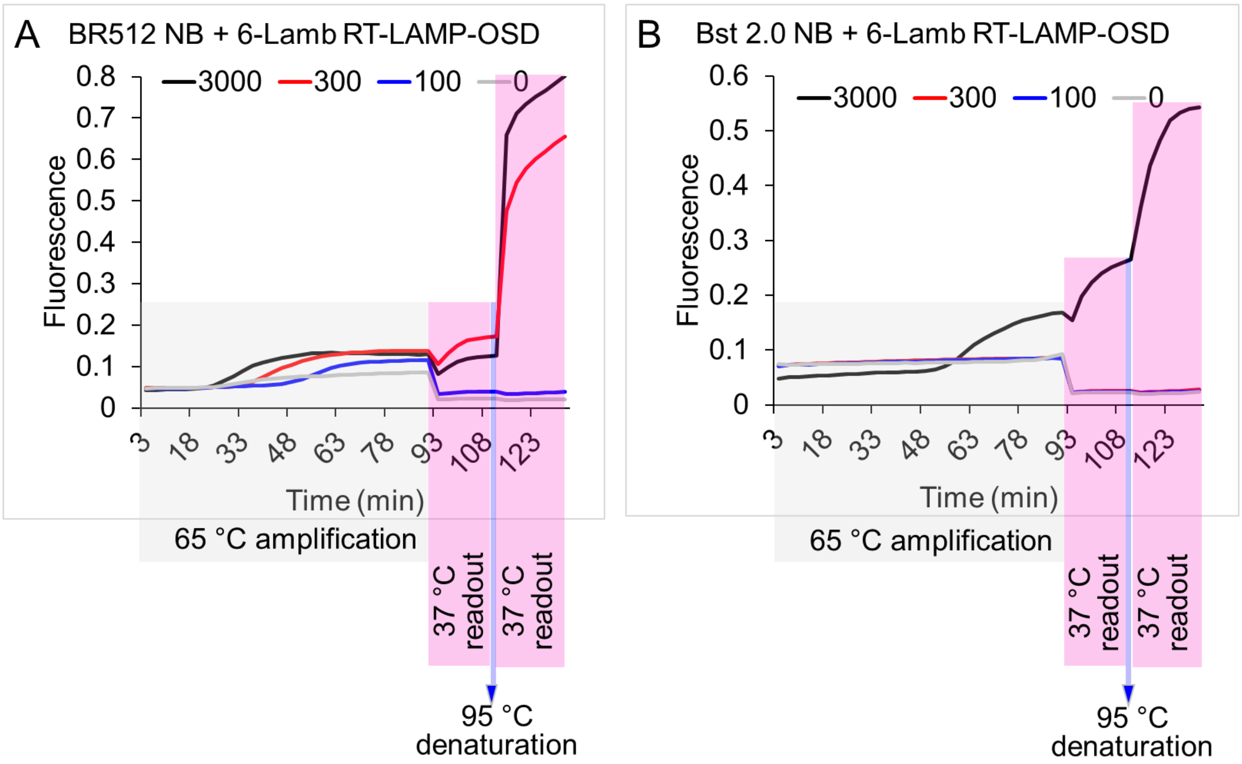


**Supplementary Figure 6. Comparison of Br512 and Bst2.0 SARS-CoV-2 multiplex RT-LAMP-OSD assays.** Duplicate multiplex RT-LAMP-OSD assays containing primers and OSD probes for both NB and 6-Lamb assays were executed with either 20 pm of Br512 in G6D buffer (A) or 16 units of Bst 2.0 in isothermal buffer (NEB) (B). Assays were seeded with indicated copies of SARS-CoV-2 genomic RNA and amplification kinetics at 65 °C observed in real-time by measuring OSD fluorescence are depicted in gray shaded boxes as black traces (3000 copies), red traces (300 copies), blue traces (100 copies), and gray traces (0 copies). Post-amplification OSD signal measured at 37 °C before and after a 1 min DNA denaturation step at 95 °C are depicted in the pink shaded regions.

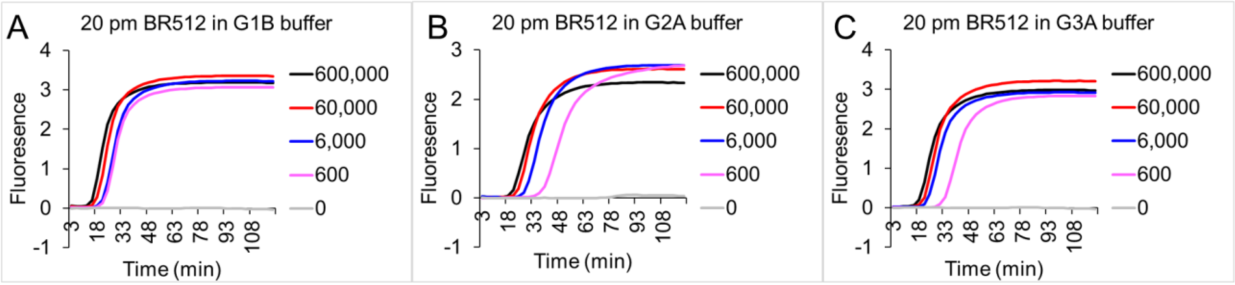


**Supplementary Figure 7. Comparison of Br512 activity in different LAMP-OSD assay buffers.** LAMP-OSD assays for human *gapd* gene were operated using Br512 in indicated reaction buffers. Amplification curves observed in real-time by measuring OSD fluorescence at 65 °C in reactions seeded with 600,000 (black traces), 60,000 (red traces), 6,000 (blue traces), 600 (pink traces), and 0 (gray traces) copies of *gapd* plasmid templates are depicted.

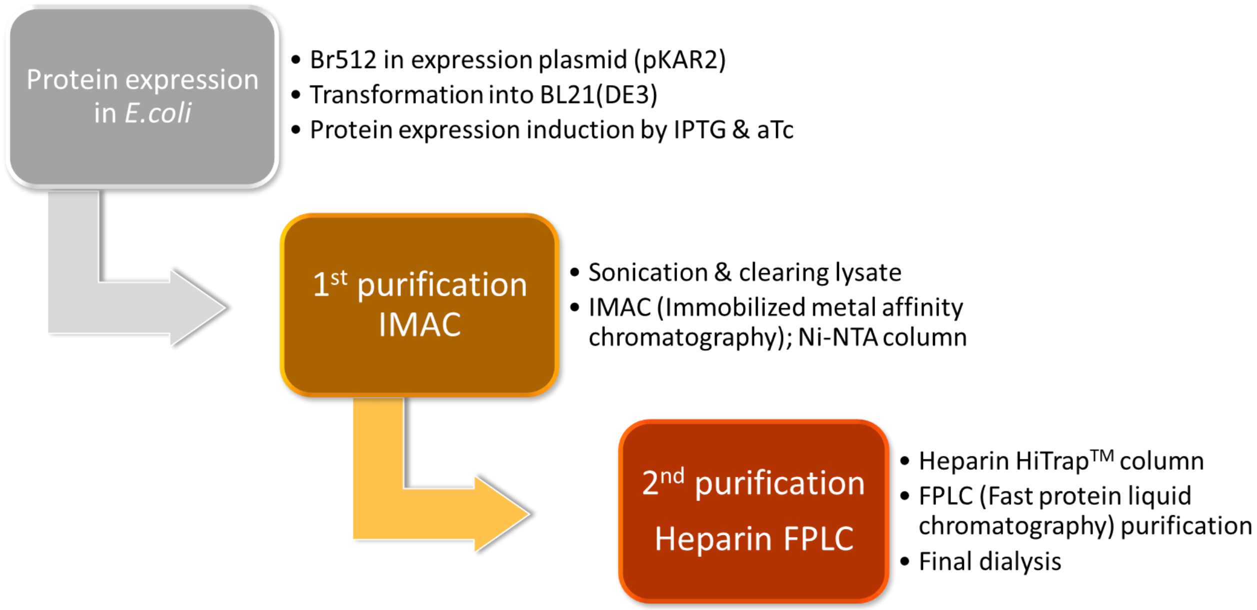


**Supplementary Figure 8. A flowchart of simple two-step Br512 purification.** Simple two-step purification procedures are shown in the flowchart. *E. coli* BL21(DE3) cell expressed Br512 was initially purified with Ni-NTA based immobilized metal affinity chromatography (IMAC), and further purified with heparin column based FPLC. A detailed purification protocol is described in the materials and methods section.

**Supplementary Table 1. Oligonucleotide and template sequences used in the study**

| Name | Sequence | Use |
|---|---|---|
| gapdLAMP.F3 | GCCACCCAGAAGACTGTG | gapd LAMP-OSD |
| gapdLAMP.B3 | TGGCAGGTTTTTCTAGACGG | |
| gapdLAMP.FIP | CGCCAGTAGAGGCAGGGATGAGGGAAACTGTGGCGTGAT | |
| gapdLAMP.BIP | GGTCATCCCTGAGCTGAACGGTCAGGTCCACCACTGACAC | |
| gapdLAMP.LR | TGTTCTGGAGAGCCCCGCGGCC | |
| gapdOSD.F | /56-FAM/CTCACTGGCATGGCCTTCCGTGTCCCCACTGCCAAC/3InvdT/ | |
| gapdOSD.Q | GGACACGGAAGGCCATGCCAGTGAG/3IABkFQ/ | |
| gapd template | CTAGTAACGGCCGCCAGTGTGCTGGAATTCCCACAGTCCATGCCATCAC TGCCACCCAGAAGACTGTGGATGGCCCCTCCGGGAAACTGTGGCGTGA TGGCCGCGGGGCTCTCCAGAACATCATCCCTGCCTCTACTGGCGCTGC CAAGGCTGTGGGCAAGGTCATCCCTGAGCTGAACGGGAAGCTCACTGG CATGGCCTTCCGTGTCCCCACTGCCAACGTGTCAGTGGTGGACCTGAC CTGCCGTCTAGAAAAACCTGCCAAATATGATGACATCAAGAAGGTGGTG AAGCAGGCGTCGGAGGGCCCCCTCAAGGGCATCCTGGGCTACACTGA GCACCAGGTGGTCTCCTCTGACTTCAACAGCGACACCCACTCCTCCACC TTTGACGCTGGGGCTGGCATTGCCCTCAACGACCACTTTGTCAAGCTCA TTTCCTGGAATTCTGCAGATATCCATCACACTGGCGGCCGCTCGAGC | |
| NB-F3 | ACCGAAGAGCTACCAGACG | NB RT-LAMP-OSD |
| NB-B3 | TGCAGCATTGTTAGCAGGAT | |
| NB-FIP | TCTGGCCCAGTTCCTAGGTAGTTCGTGGTGGTGACGGTAA | |
| NB-BIP | AGACGGCATCATATGGGTTGCACGGGTGCCAATGTGATCT | |
| NB-LB | ACTGAGGGAGCCTTGAATACA | |
| NB-OSD-FAM | /56-FAM/CCGAATGAAAGATCTCAGTCCAAGATGGTATTTCT/3InvdT/ | |
| NB-OSD-Q | TCTTGGACTGAGATCTTTCATTCGG/3IABkFQ/ | |
| Lamb-F3 | TCCAGATGAGGATGAAGAAGA | 6-Lamb RT-LAMP-OSD |
| Lamb-B3 | AGTCTGAACAACTGGTGTAAG | |
| Lamb-FIP | AGAGCAGCAGAAGTGGCACAGGTGATTGTGAAGAAGAAGAG | |
| Lamb-BIP | TCAACCTGAAGAAGAGCAAGAACTGATTGTCCTCACTGCC | |
| Lamb-LF | CTCATATTGAGTTGATGGCTCA | |
| Lamb-LB | ACAAACTGTTGGTCAACAAGAC | |
| Lamb-OSD-FAM | GTATGGTACTGAAGATGATTACCAAGGTAAACCTTTGGAATTTGGAC/36-FAM/ | |
| Lamb-OSD-Q | /5IABkFQ/GTCCAAATTCCAAAGGTTTACCTTGGTAATCATCTC/3InvdT/ | |
| Tholoth-F3 | TGCTTCAGTCAGCTGATG | Tholoth RT-LAMP-OSD |
| Tholoth-B3 | TTAAATTGTCATCTTCGTCCTT | |
| Tholoth-FIP | TCAGTACTAGTGCCTGTGCCCACAATCGTTTTTAAACGGGT | |
| Tholoth-BIP | TCGTATACAGGGCTTTTGACATCTATCTTGGAAGCGACAACAA | |
| Tholoth-LB | GTAGCTGGTTTTGCTAAATTCC | |
| Tholoth-OSD-FAM | /56-FAM/ACAGGTGTAAGTGCAGCCCGTCTTACACCGTGC/3InvdT/ | |
| Tholoth-OSD-Q | GACGGGCTGCACTTACACCTGT/3IABkFQ/ | |

**Supplementary Table 2. Full sequence of pKAR2-Br512; 6218 bp.** Br512 is highlighted.

Detailed annotation of Br512
4285-4311 : 8x His
4312-4452 : HP47
4453-4476 : 2(GS)3(A)P
4476-6216 : Bst-LF

ATCCTAAGCTTAATTTAGCATAACCCCTTGGGGCCTCTAAACGGGTCTTGAGGGGTTTTTT
GAGCTGAGCTTGGACTCCTGTTGATAGATCCGGCCGGTAATGACCTCAGAACTCCATCTG
CCTAATGGAGTGATTCTAGCCGGTCGTTACACGTGGAACGGGAACTGCCAGACATCAAATA
AAACAAAAGGCTCAGTCGGAAGACTGGGCCTTTTGTTTTATCTGTTGTTTGTCGGTGAACA
CTCTCCCGGAAAACTCACGTTAAGGGATTTTGGTCATGAGATTATCAAAAAGGATCTTCACC
TAGATCCTTTTAAACTAGTGAAGTTACCATCACGGAAAAAGGTTATGCTGCTTTTAAGACCC
ACTTTCACATTTAAGTTGTTTTTCTAATCCGCAAATGATCAATTCAAGGCCGAATAAGAAGG
CTGGCTCTGCACCTTGGTGATCAAATAATTCGATAGCTTGTCGTAATAATGGCGGCATACT
ATCAGTAGTAGGTGTTTCCCTTTCTTCTTTAGCGACTTGATGCTCTTGATCTTCCAATACGC
AACCTAAAGTAAAATGCCCCACTGCACTGAGTGCATATAATGCATTCTCTAGTGAAAAACCT
TGTTGGCATAAAAAGGCTAATTGATTTTCGAGAGTTTCATACTGTTTTTCTGTAGGCCGTGT
ACCTAAATGTACTTTTGCTCCATCGCGATGACTTAGTAAAGCACATCTAAAACTTTTAGCGT
TATTACGTAAAAAATCTTGCCAGCTTTCCCCTTCTAAAGGGCAAAAGTGAGTATGGTGCCTA
TCTAACATCTCAATGGCTAAGGCGTCGAGCAAAGCCCGCTTATTTTTTACATGCCAATACAA
TGTAGGCTGCTCTACACCTAGCTTCTGGGCGAGTTTACGGGTTGTTAAACCTTCGATTCCG
ACCTCATTAAGCAGCTCTAATGCGCTGTTAATCACTTTACTTTTATCTAAACGAGACATCATT
AACCTCCTCAAGAGGATCGAATAGTTATTACCAATGCTTAATCAGTGAGGCACCTATCTCAG
CGATCTGTCTATTTCGTTCATCCATAGTTGCCTGACTCCCCGTCGTGTAGATAACTACGATA
CGGGAGGGCTTACCATCTGGCCCCAGTGCTGCAATGATACCGCGAGAGCCACGCTCACC
GGCTCCAGATTTATCAGCAATAAACCAGCCAGCCGGAAGGGCCGAGCGCAGAAGTGGTCC
TGCAACTTTATCCGCCTCCATCCAGTCTATCAATTGTTGCCGGGAAGCTAGAGTAAGTAGT
TCGCCAGTTAATAGTTTGCGCAACGTTGTTGCCATTGCTACAGGCATCGTGGTGTCACGCT
CGTCGTTTGGTATGGCTTCATTCAGCTCCGGTTCCCAACGATCAAGGCGAGTTACATGATC
CCCCATGTTGTGCAAAAAAGCGGTTAGCTCCTTCGGTCCTCCGATCGTTGTCAGAAGTAAG
TTGGCCGCAGTGTTATCACTCATGGTTATGGCAGCACTGCATAATTCTCTTACTGTCATGCC
ATCCGTAAGATGCTTTTCTGTGACTGGTGAGTACTCAACCAAGTCATTCTGAGAATAGTGTA
TGCGGCGACCGAGTTGCTCTTGCCCGGCGTCAATACGGGATAATACCGCGCCACATAGCA
GAACTTTAAAAGTGCTCATCATTGGAAAACGTTCTTCGGGGCGAAAACTCTCAAGGATCTTA
CCGCTGTTGAGATCCAGTTCGATGTAACCCACTCGTGCACCCAACTGATCTTCAGCATCTT
TTACTTTCACCAGCGTTTCTGGGTGAGCAAAAACAGGAAGGCAAAATGCCGCAAAAAAGGG
AATAAGGGCGACACGGAAATGTTGAATACTCATACTCTTCCTTTTTCAATATTATTGAAGCA
TTTATCAGGGTTATTGTCTCATGAGCGGATACATATTTGAATGTATTTAGAAAAATTTTTTAA
GGCAGTTATTGGTGCCGCTTAAACGCCTGGGGTAATGACTCTCTAGCTTGAGGCATCAAAT
AAAACGAAAGGCTCAGTCGAAAGACTGGGCCTTTCGTTTTATCTGTTGTTTGTCGGTGAAC
GCTCTCCTGAGTAGGACAAATCCGCCCTCTAGATTACGTGCAGTCGATGATAAGCTGTCAA
ACGGAATTTCGGGCAGCGTTGGGTCCTGGCCACGGGTGCGCCGGTGTGAAATACCGCAC
AGATGCGTAAGGAGAAAATACCGCATCAGGCGCTCTTCCGCTTCCTCGCTCACTGACTCG
CTGCGCTCGGTCGTTCGGCTGCGGCGAGCGGTATCAGCTCACTCAAAGGCGGTAATACG
GTTATCCACAGAATCAGGGGATAACGCAGGAAAGAACATGTGAGCAAAAGGCCAGCAAAA
GGCCAGGAACCGTAAAAAGGCCGCGTTGCTGGCGTTTTTCCATAGGCTCCGCCCCCCTGA
CGAGCATCACAAAAATCGACGCTCAAGTCAGAGGTGGCGAAACCCGACAGGACTATAAAG
ATACCAGGCGTTTCCCCCTGGAAGCTCCCTCGTGCGCTCTCCTGTTCCGACCCTGCCGCT
TACCGGATACCTGTCCGCCTTTCTCCCTTCGGGAAGCGTGGCGCTTTCTCATAGCTCACGC

TGTAGGTATCTCAGTTCGGTGTAGGTCGTTCGCTCCAAGCTGGGCTGTGTGCACGAACCC
CCCGTTCAGCCCGACCGCTGCGCCTTATCCGGTAACTATCGTCTTGAGTCCAACCCGGTA
AGACACGACTTATCGCCACTGGCAGCAGCCACTGGTAACAGGATTAGCAGAGCGAGGTAT
GTAGGCGGTGCTACAGAGTTCTTGAAGTGGTGGCCTAACTACGGCTACACTAGAAGGACA
GTATTTGGTATCTGCGCTCTGCTGAAGCCAGTTACCTTCGGAAAAAGAGTTGGTAGCTCTT
GATCCGGCAAACAAACCACCGCTGGTAGCGGTGGTTTTTTTGTTTGCAAGCAGCAGATTAC
GCGCAGAAAAAAAGGATCTCAAGAAGATCCTTTGATCTTTTCTACGGGGTCTGACGCTCAG
TGGAACGAAAACTCACGTTAAGGGATTTTGGTCATGGAATTAATTCTTAGAAAAACTCATCG
AGCATCAAATGAAACTGCAATTTATTCATATCAGGATTATCAATACCATATTTTTGAAAAAGC
CGTTTCTGTAATGAAGGAGAAAACTCACCGAGGCAGTTCCATAGGATGGCAAGATCCTGGT
ATCGGTCTGCGATTCCGACTCGTCCAACATCAATACAACCTATTAATTTCCCCTCGTCAAAA
ATAAGGTTATCAAGTGAGAAATCACCATGAGTGACGACTGAATCCGGTGAGAATGGCAAAA
GTTTATGCATTTCTTTCCAGACTTGTTCAACAGGCCAGCCATTACGCTCGTCATCAAAATCA
CTCGCATCAACCAAACCGTTATTCATTCGTGATTGCGCCTGAGCGAAGACGAAATACGCGA
TCGCTGTTAAAAGGACAATTACAAACAGGAATCGAATGCAACCGGCGCAGGAACACTGCC
AGCGCATCAACAATATTTTCACCTGAATCAGGATATTCTTCTAATACCTGGAATGCTGTTTT
CCCGGGGATCGCAGTGGTGAGTAACCATGCATCATCAGGAGTACGGATAAAATGCTTGAT
GGTCGGAAGAGGCATAAATTCCGTCAGCCAGTTTAGTCTGACCATCTCATCTGTAACATCA
TTGGCAACGCTACCTTTGCCATGTTTCAGAAACAACTCTGGCGCATCGGGCTTCCCATACA
ATCGATAGATTGTCGCACCTGATTGCCCGACATTATCGCGAGCCCATTTATACCCATATAAA
TCAGCATCCATGTTGGAATTTAATCGCGGCCTAGAGCAAGACGTTTCCCGTTGAATATGGC
TCATAACACCCCTTGTATTACTGTTTATGTAAGCAGACAGTTTTATTGTGTAATCGTTAATCC
GCAAATAACGTAAAAACCCGCTTCGGCGGGTTTTTTTATGGGGGGAGTTTAGGGAAAGAG
CATTTGTCATCATGACCATGACATTAACCTATAAAAATAGGCGTATCACGAGGCCCTTTCCC
TAGGGTCTTCACACTCTATCATTGATAGAGTTAATACGACTCACTATAGGGTCCCTATCAGT
GATAGAGAGAATTCGTACTGAGCACAGCTGTCACCGGATGTGCTTTCCGGTCTGATGAGTC
CGTGAGGACGAAACAGCCTCTACAAATAATTTTGTTTAAACTAGTTAGATAAGGAGGTTACA
TATGCACCATCATCACGGTCATCACCACCATCCGCGTGGTGTTGACCCGAGCCGTAAGGA
GAACCACCTGTCTGACGAAGACTTCAAGGCGGTGTTCGGTATGACCCGTTCTGCGTTCGC
GAACCTGCCGCTGTGGAAACAACAGAACCTGAAGAAGGAGAAAGGTCTGTTCGGTTCTGG
AAGCGCAGCAGCACCTAAGATGGCATTCACATTGGCCGATCGTGTCACCGAAGAGATGCT
GGCAGACAAGGCAGCCTTGGTCGTGGAGGTAGTTGAGGAGAACTATCACGACGCACCGAT
TGTTGGAATCGCCGTGGTCAATGAACATGGTCGCTTCTTCTTGCGCCCTGAGACTGCGTTG
GCCGACCCACAATTCGTGGCCTGGTTAGGAGATGAAACGAAGAAGAAGTCAATGTTCGAC
AGCAAACGCGCAGCCGTAGCTCTGAAGTGGAAAGGAATTGAGCTGTGTGGTGTGAGTTTC
GACCTTCTCTTAGCAGCGTACTTGCTTGATCCCGCTCAAGGCGTCGACGACGTGGCAGCC
GCTGCCAAGATGAAGCAATATGAAGCGGTGCGTCCGGATGAGGCTGTGTACGGGAAGGG
AGCTAAACGCGCGGTGCCTGATGAACCCGTGCTTGCTGAGCACTTGGTACGCAAGGCTGC
GGCTATCTGGGAGCTGGAGCGTCCCTTCCTGGATGAGTTGCGTCGCAACGAGCAGGACC
GCCTGCTTGTAGAGTTAGAACAGCCTCTTAGCTCTATTCTTGCCGAGATGGAGTTCGCTGG
TGTCAAAGTAGATACCAAGCGCCTTGAGCAAATGGGTAAGGAGTTGGCTGAACAACTGGG
CACAGTGGAACAGCGTATCTACGAACTGGCCGGTCAGGAGTTCAACATCAACAGCCCCAA
GCAGCTGGGAGTGATCCTGTTCGAGAAGTTGCAGCTGCCAGTATTGAAGAAGACTAAGAC
TGGCTACAGTACCTCGGCTGACGTACTGGAGAAGCTGGCTCCTTACCATGAGATCGTGGA
GAACATCTTGCACTACCGCCAGCTGGGCAAGCTGCAGTCTACCTACATTGAGGGTCTGTTA
AAGGTCGTGCGTCCAGACACGAAGAAGGTGCATACGATCTTCAATCAGGCGCTGACCCAA
ACTGGTCGTTTGTCGTCCACAGAGCCCAATCTTCAGAATATCCCTATTCGTCTTGAGGAAG
GCCGCAAGATTCGCCAGGCCTTCGTTCCTTCGGAATCGGACTGGCTGATCTTCGCAGCAG
ATTACTCACAGATCGAGCTTCGCGTGTTGGCACATATCGCGGAGGATGACAACTTAATGGA
GGCGTTCCGCCGCGATCTGGATATCCATACTAAGACCGCGATGGATATCTTCCAAGTGTCA
GAAGACGAGGTAACACCGAACATGCGACGCCAGGCGAAAGCGGTTAACTTCGGCATCGTC

TACGGCATCAGCGACTATGGCCTGGCCCAGAACTTGAACATCAGCCGCAAGGAGGCAGCC
GAGTTCATCGAGCGCTACTTCGAGAGTTTCCCAGGTGTGAAGCGTTATATGGAGAATATCG
TACAAGAGGCGAAGCAGAAAGGCTACGTGACCACGCTGTTACACCGTCGTCGCTACCTTC
CTGATATCACTAGCCGTAACTTCAATGTACGTTCCTTCGCCGAACGCATGGCGATGAATAC
CCCCATCCAGGGGTCAGCTGCAGATATCATCAAGAAAGCTATGATCGACTTAAACGCTCGT
CTGAAGGAAGAACGCTTACAGGCGCACCTCTTACTGCAAGTCCATGACGAATTGATCCTTG
AGGCGCCCAAGGAAGAGATGGAGCGTCTTTGCCGTCTGGTGCCGGAAGTAATGGAACAG
GCCGTCACGCTGCGCGTACCTCTGAAAGTCGATTACCACTACGGCTCCACCTGGTATGAC
GCCAAGTAAGG
